# Epidemiology and precision of SARS-CoV-2 detection following lockdown and relaxation measures

**DOI:** 10.1101/2020.09.22.20198697

**Authors:** Karoline Leuzinger, Rainer Gosert, Kirstine K. Søgaard, Klaudia Naegele, Julia Bielicki, Tim Roloff, Roland Bingisser, Christian H. Nickel, Nina Khanna, Sarah Tschudin Sutter, Andreas F. Widmer, Katharina Rentsch, Hans Pargger, Martin Siegemund, Daiana Stolz, Michael Tamm, Stefano Bassetti, Michael Osthoff, Manuel Battegay, Adrian Egli, Hans H. Hirsch

**Affiliations:** Clinical Virology, Laboratory Medicine, University Hospital Basel, Basel, Switzerland; Transplantation & Clinical Virology, Department Biomedicine, University of Basel, Basel, Switzerland; Applied Microbiology Research, Laboratory Medicine, Department Biomedicine, University of Basel, Basel, Switzerland; Clinical Bacteriology and Mycology, Laboratory Medicine, University Hospital Basel, Basel, Switzerland; Pediatric Infectious Diseases & Hospital Epidemiology, University Children Hospital Basel, Basel, Switzerland; Emergency Medicine, University Hospital Basel, Basel, Switzerland; Infectious Diseases & Hospital Epidemiology, University Hospital Basel; Clinical Chemistry, Laboratory Medicine, University Hospital Basel, Basel, Switzerland; Intensive Care Unit, University Hospital Basel, Basel, Switzerland; Clinic of Pneumology and Pulmonary Cell Research, University Hospital Basel, Basel, Switzerland; Internal Medicine, University Hospital Basel, Basel, Switzerland

**Keywords:** Severe acute respiratory syndrome coronavirus 2, COVID-19, Basel-SCoV2-S111bp, Basel-SCoV2-ORF8-98bp, bronchoalveolar lavage, tracheal secretion, NAAT, QNAT, PCR

## Abstract

**Introduction:** SARS-CoV-2-detection is critical for clinical and epidemiological assessment of the ongoing CoVID-19 pandemic.

**Aim:** To cross-validate manual and automated high-throughput (Roche-cobas®6800-Target1/Target2) testing for SARS-CoV-2-RNA, to describe detection rates following lockdown and relaxation, and to evaluate SARS-CoV-2-loads in different specimens.

**Method:** The validation cohort prospectively compared Basel-S-gene, Roche-E-gene, and Roche-cobas®6800-Target1/Target2 in 1344 naso-oropharyngeal swabs (NOPS) taken in calendar week 13 using Basel-ORF8-gene-assay for confirmation. Follow-up-cohort-1 and -2 comprised 12363 and 10207 NOPS taken over 10 weeks until calendar week 24 and 34, respectively. SARS-CoV-2-loads were compared in follow-up NOPS, lower respiratory fluids, and plasma.

**Results:** Concordant results were obtained in 1308 cases (97%) including 97 (9%) SARS-CoV-2-positives showing high quantitative correlations (Spearman r>0.95; p<0.001) for all assays. Discordant samples (N=36) had significantly lower SARS-CoV-2-loads (p<0.001). Following lockdown, weekly detection rates declined to <1% reducing single-test positive predictive values from 99.3% to 85.1%. Following relaxation, rates flared up to 4% with similarly high SARS-CoV-2-loads, but patients were significantly younger than during lockdown (34 vs 52 years, p<0.001). SARS-CoV-2-loads in follow-up NOPS declined by 3log10 copies/mL within 10 days post-diagnosis (p<0.001). SARS-CoV-2-loads in NOPS correlated weakly with those in time-matched lower respiratory fluids and plasma, but remained detectable in 14 and 7 cases of NOPS with undetectable SARS-CoV-2, respectively.

**Conclusion:** Evaluated manual and automated assays are highly concordant and correlate quantitatively. Following successful lockdown, declining positive predictive values require dual-target-assays for clinical and epidemiologic assessment. Confirmatory and quantitative follow-up testing should be considered within <5 days, using lower respiratory fluids in symptomatic patients with SARS-CoV-2-negative NOPS.

## Introduction

SARS-CoV-2 is the cause of the coronavirus infectious disease called CoVID-19, which surfaced in late 2019 as a novel global public health emergency [1, 2]. In the middle of August 2020, Europe alone was confronted with 3.7 million reported SARS-CoV-2 infections and >210’000 deaths and the dire prospects of exceeding 4 million by the end of summer [3]. Given the continuing global transmission, the unavailability of effective and safe vaccines or antiviral agents for prophylaxis or treatment, many countries have implemented targeted lockdown- and containment strategies as recommended by the WHO [4]. To this end, prompt testing of symptomatic individuals [2, 5] as well as follow-up testing of exposed and recovering patients potentially shedding SARS-CoV-2 are important for case identification, contact tracing, infection control, and clinical patient management [6-8]. Detecting viral RNA in respiratory fluids by reverse transcription–quantitative nucleic acid testing (RT-QNAT) is currently the gold standard for the specific diagnosis of SARS-CoV-2 replication and CoVID-19 [9]. The rapid availability of viral genome sequences in January 2020 facilitated the development of RT-QNAT protocols for SARS-CoV-2 detection [10-12], followed by a range of commercial SARS-CoV-2 assays in the subsequent months [12-15]. In our center, we applied a prospective parallel testing strategy using the laboratory-developed Basel-S-gene RT-QNAT assay together with a commercial assay for rapid delivery of confirmed SARS-CoV-2 results in the early pandemic phase until March 2020 [11]. To meet the increasing demand of SARS-CoV-2 tests, and to reduce the hands-on time in the laboratory, we prospectively validated a fully automated SARS-CoV-2-assay. Here, we report on the prospective comparison of our established approach with the automated high-throughput Roche-cobas®6800-Target1/Target2 using nasopharyngeal-oropharyngeal swabs (NOPS). As comprehensive real-life data are scarce, we analyze the epidemiology of SARS-CoV-2-detection following lockdown and relaxation measures and investigate the quantitative relationship of the different assays and explore SARS-CoV-2-loads in follow-up NOPS and in time-matched lower respiratory fluids, and in plasma samples.

## Methods

### Study periods and clinical samples

The prospective validation cohort consisted of 1344 naso-oropharyngeal swabs (NOPS) submitted from 30 March to 3 April 2020 in calendar week 13, which were analyzed in parallel with the Basel-SCoV2-S-111bp targeting the S-gene of SARS-CoV-2, the commercial Roche-E-gene RT-QNAT and the cobas® SARS-CoV-2 assay (Roche-Cobas-Target1/Target2). The Basel-SCoV2-ORF8-98bp corresponding to the previously described as Basel-N-gene assay was used as confirmatory assay for discordant NOPS [11]. Subsequently, 12363 NOPS were analyzed with the Roche-Cobas-Target1/Target2 assay from calendar week 14 to 24 (follow-up cohort-1) and 10207 NOPS from calendar week 24 to 34 (follow-up cohort-2) using Basel-S-gene for discordant Roche-Cobas-Target1 and - Target2 results. For NOPS sampling, nasopharyngeal and oropharyngeal sites were swabbed separately and collected together in one universal transport medium tube (UTM) as described [11]. Basel-S-gene RT-QNAT was used for SARS-CoV-2 genome quantification in 936 follow-up NOPS from 261 patients with a positive Roche-Cobas-Target1/Target2 screening result, in 95 NOPS and time-matched lower respiratory fluids (tracheal aspirates, bronchial-alveolar lavage, or sputum) or in 259 NOPS and time-matched plasma samples from COVID-19 patients.

### Total nucleic acid extraction and reverse transcription quantitative nucleic acid test

Total nucleic acids (TNAs) were extracted from the UTMs lower respiratory fluids and plasma using the DNA and viral NA small volume kit on the MagNA Pure 96 system (Roche Diagnostics, Rotkreuz, Switzerland) or the Abbott sample preparation system reagent kit using the Abbott m2000 Realtime System (Abbott, Baar, Switzerland). SARS-CoV-2 with the commercial Roche-E-gene assay followed the guidelines provided by the manufacturer using the CFX96 RT-PCR system (Bio-Rad Laboratories, Cressier, Switzerland). Cycle threshold numbers were obtained for quantitative comparisons and copy number derived from standard curves run in parallel. SARS-CoV-2 loads were determined in NOPS, lower respiratory fluids, or in plasma using Basel-SCoV2-S-111bp and Basel-SCoV2-ORF8-98bp [11].

### Cobas® SARS-CoV-2 assay

NOPS were automatically processed by the cobas® 6800 system, TNA from patient samples were extracted together with the RNA internal control. As described by the manufacture, sequences of the open reading frame (ORF)-1 of the non-structural region served as SARS-CoV-2-specific Roche-Cobas-Target1, while conserved regions in the structural E-gene served as Roche-Cobas-Target2.

### Assessing genetic variation in SARS-CoV-2 genome sequences

SARS-CoV-2 high coverage, complete genome sequences (N=25’168) were downloaded from the NCBI-GenBank and GISAID database (https://www.gisaid.org/; accessed on 23 July 2020). The frequency of single nucleotide polymorphisms (SNPs) was analyzed in complete SARS-CoV-2 genome sequences, in the spike glycoprotein (S)-gene, the envelope (E)-gene, and the ORF8-gene sequences using the basic variant detection tool of the CLC Genomic Workbench software (version 12; QIAGEN, Hilden, Germany), and a viral reference genome (acc. no. NC_045512) as described previously [16].

### Biofire Filmarray respiratory panel

Twenty NOPS testing positive for human coronavirus (HCoV)-HKU1 (Betacoronavirus, Embecovirus), HCoV-OC43 (Betacoronavirus, Embecovirus), HCoV-229E (Alphacoronavirus, Duvinacovirus) or HCoV-NL63 (Alphacoronavirus, Setracovirus) in the multiplex NAT respiratory panel (Torch system; Biofire Filmarray respiratory 2.0 panel, bioMérieux) were analyzed for target specificity using Basel-SCoV2-S-111bp, Basel-SCoV2-ORF8-98bp, Roche-E-gene, and the Roche-Cobas-Target1/Target2 assays.

### Statistical analysis

All statistical data analysis was done in R (version 3.6.1; https://cran.r-project.org), and Prism (version 8; Graphpad Software, CA, USA) was used for data visualization. Statistical comparison of non-parametric data was done using Mann-Whitney U test. Spearman’s rank correlation and Bland-Altman analysis was used to assess the relationship between cycles thresholds of the different SARS-CoV-2 assays reflecting SARS-CoV-2 RNA loads. Regression analysis was performed to assess the decrease in SARS-CoV-2 load after diagnosis.

### Ethics Statement

The study was conducted according to good laboratory practice and in accordance with the Declaration of Helsinki and national and institutional standards for laboratory quality control and was approved by the Ethical Committee of North-western and Central Switzerland (EKNZ 2020-00769).

## Results

### Cross-validation of manual and automated SARS-CoV-2 RNA detection assays

To validate the automated high-throughput Roche-Cobas-SARS-CoV-2-Target1/Target2 assay on the cobas®6800 platform, 1344 NOPS taken in calendar week 13 were prospectively tested in parallel with the previously validated Basel-S-gene and Roche-E-gene assays [11]. The samples had been submitted from 1255 (93%) symptomatic adult and 89 (7%) pediatric patients (**Table 1**). Concordant results were obtained in 1308 (97%) samples, consisting of 97 (7%) positive and 1211 (90%) negative cases (**Supplementary Figure S1**). Discordant results were obtained in 36 (3%) cases, mostly consisting of Basel-S-gene-positive cases (N=14) or Basel-S-gene and Roche-Cobas-Target1 and Target2-positive cases (N=12). In 20/36 (56%) cases, SARS-CoV-2 detection was independently confirmed by the Basel-ORF8-gene RT-QNAT consisting of 9/14 and 11/12 of the 36 discordant cases (**Supplementary Figure S1**). Accordingly, the Roche-Cobas-Target1 assay had a sensitivity of 92.3% (95% CI: 85.9% - 96.4%) and a specificity of 99.8% (95% CI: 99.4% - 100%), while the Roche-Cobas-Target2 assay had a sensitivity of 92.3% (95% CI: 85.9% - 96.4%) and a specificity of 99.2% (95% CI: 98.5% - 99.6%)(**Supplementary Table S1**).

**Table 1.**
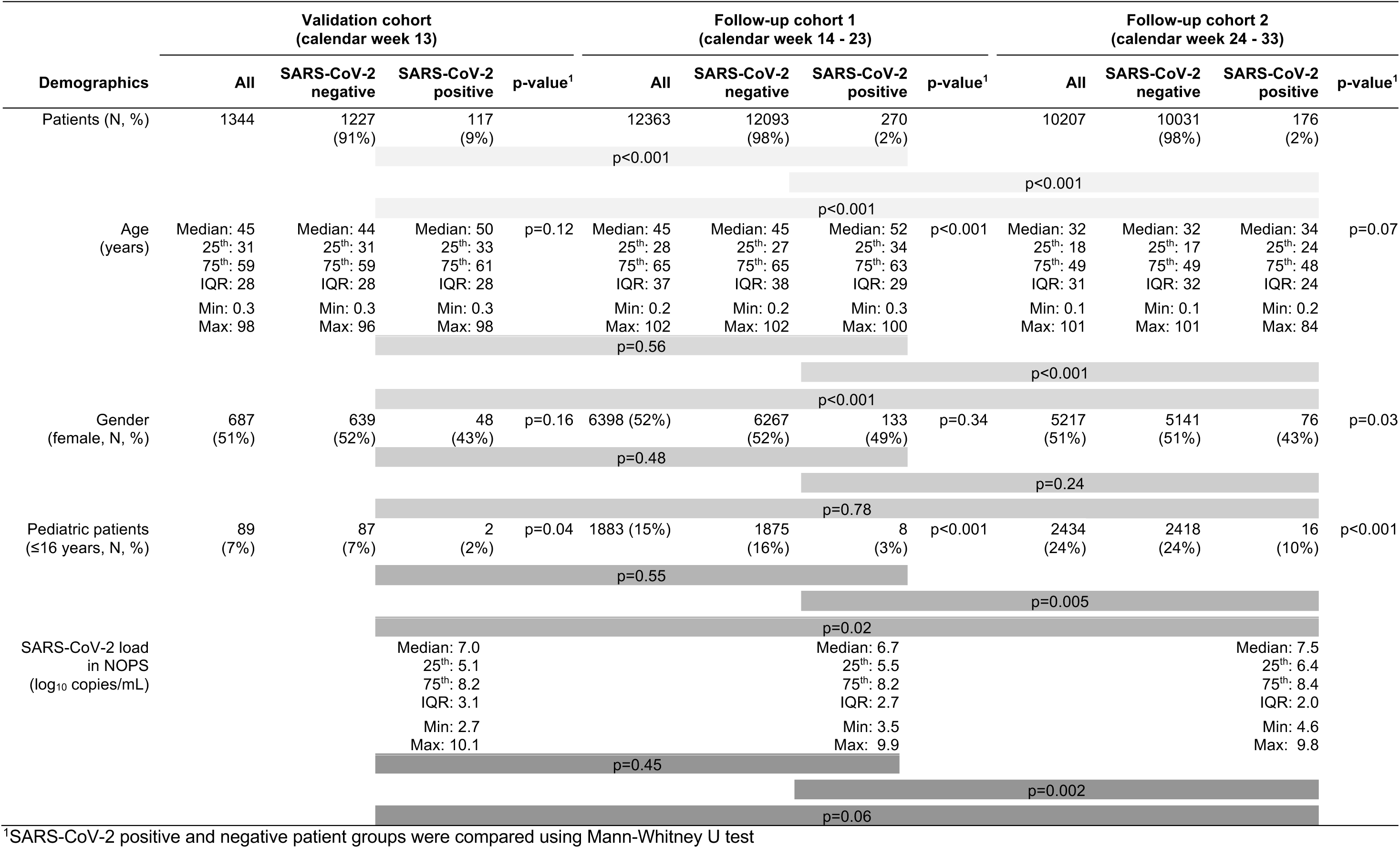
Patients’ demographics.

Importantly, the cycle threshold (Ct) values were significantly lower indicating higher viral loads for the concordant positive samples than for the discordant ones being close to the limit of detection (p<0.001; **Figure 1**).

**Figure 1.**
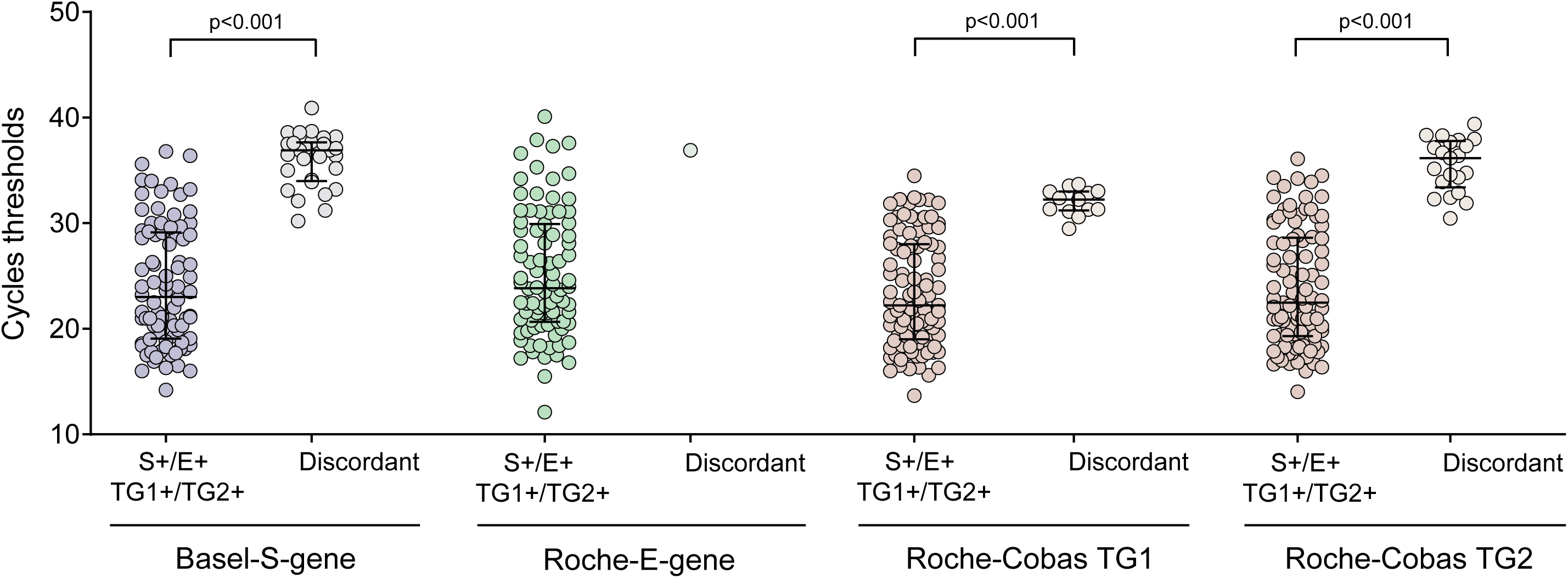
Comparison of cycles thresholds in the Basel-S-gene, Roche-E-gene and the Roche Cobas Target1/Target2 assays. NOPS were prospectively tested in parallel with the laboratory-developed Basel-S-gene, the commercial Roche-E-gene and the Roche-Cobas-Target1/Target2 assays (N=1344). Cycles thresholds of concordant positive and discordant samples are displayed (median, 25th and 75th percentiles; N=1344).

To address the diversity of SARS-CoV-2 genomes potentially impacting assay performance and discordance rate, 25’168 complete sequences were downloaded and analyzed. Overall, the SARS-CoV-2 genomes were highly conserved with only eight SNPs present at frequencies >10% within the large 30’000 nt-long RNA genome sequence (**Figure 2**). The S-gene sequence of 3’822 nt displayed high sequence conservation with only two SNPs present at frequencies >2% (A23403G at 74.1% and C23731T at 2.3%). Importantly, only a single SNP (i.e. C22432T) was detected in the probe-binding site of the Basel-S-gene RT-QNAT in 268 of 25’168 (1.1%) sequences, at a position not predicted to affect the assay performance (**Figure 2**). The ORF8-gene sequences of 366 nt showed SNPs at C27964T, T28144C and G28077C at frequencies of 3.2%, 11.7%, and 1.1%, respectively. Target sequences of the commercial assays are not publicly available precluding evaluation of the assay targets, but the overall SNP frequencies of the E-gene were below 0.3% (**Figure 2**). To specifically address the impact of circulating HCoVs, NOPS from 20 different patients testing positive for each of the four HCoV-HKU1, -NL63, -OC43, and -229E were analyzed, but showed no cross-detection by any of the five assays (**Supplementary Table 2**).

**Figure 2.**
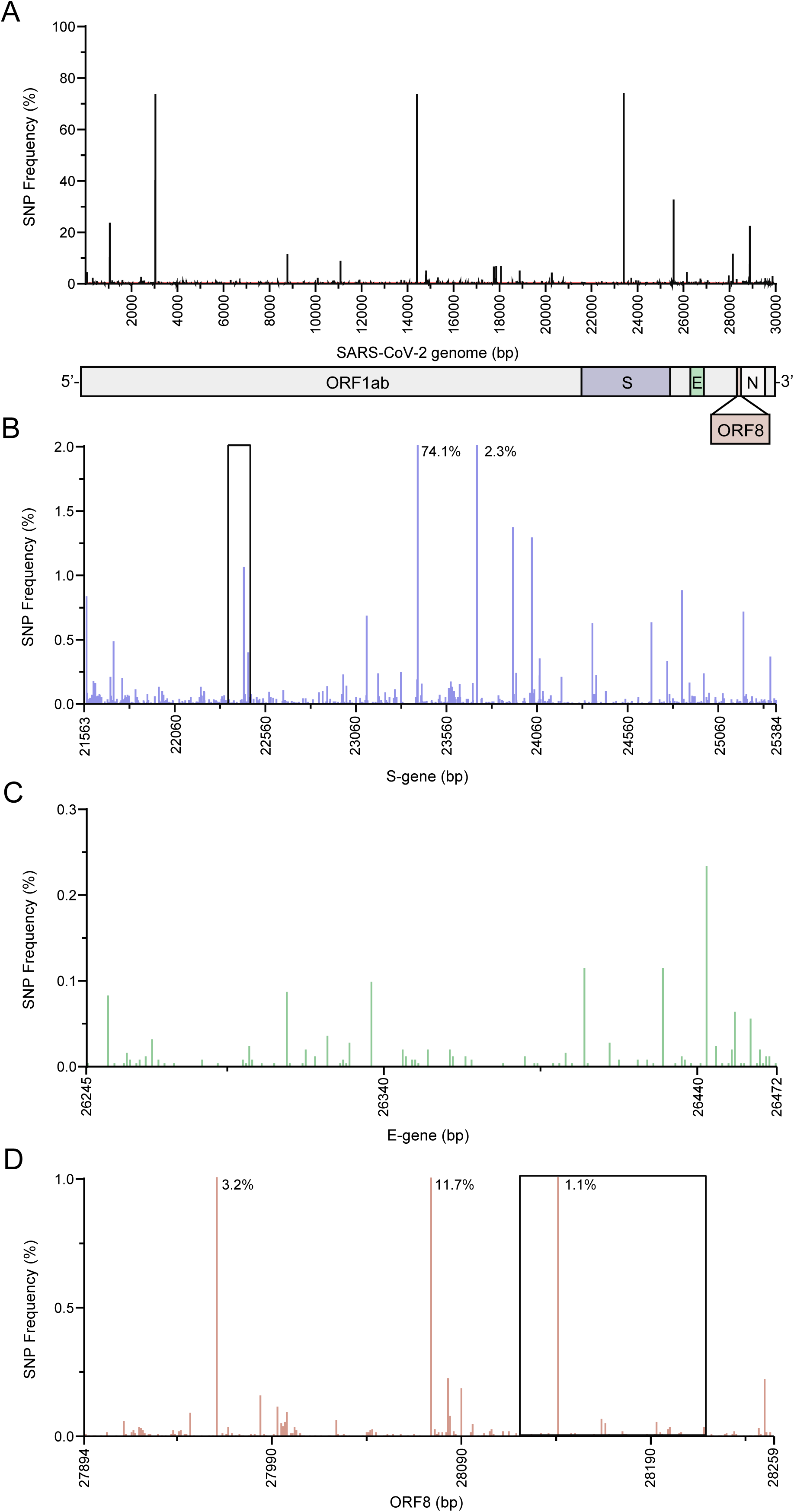
Detection of genetic variation in complete SARS-CoV-2 genome sequences, S-gene, E-gene and ORF8-gene sequences. The frequency of single nucleotide polymorphisms (SNPs) was assessed in the complete genome sequences, the S-gene, E-gene and ORF8-gene sequences using the basic variant detection tool of the CLC Genomic Workbench software (QIAGEN). A. Genetic variation in complete SARS-CoV-2 genome sequences of 29’900 nucleotides (nt) in length available in the NCBI-GenBank and GISAID database (accessed on 23 July 2020; N=25168). Schematic representation of the SARS-CoV-2 genome organization (nucleotide positions correspond to the SARS-CoV-2 reference genome [acc. no. NC_045512.2]). B. Genetic variation in the S-gene region of 3’822 nt (corresponds to nucleotide positions 21563 to 25384 in the SARS-CoV-2 reference genome [acc. no. NC_045512.2]; n=25168). Basel-S-gene QNAT target region is marked with a black frame. C. Genetic variation in the E-gene region of 228 nt (corresponds to nucleotide positions 26245 to 26472 in the SARS-CoV-2 reference genome [acc. no. NC_045512.2]; N=25168). D. Genetic variation in the ORF8-gene region of 366 nt (corresponds to nucleotide positions 27894 to 28259 in the SARS-CoV-2 reference genome [acc. no.NC_045512.2]; N=25168). Basel-ORF8-gene QNAT target region is marked with a black frame.

Taken together, the results indicated a very good performance of all assays, and suggested a stochastic distribution of SARS-CoV-2 in NOPS carrying very low viral loads as the most likely reason for discordance. Thus, the prospective validation cohort of 1344 patients comprised a total of 117 (9%) confirmed SARS-CoV-2 infections. Compared to those with a negative test result, the demographics revealed no significant differences (**Table 1**).

### SARS-CoV-2 detection rates following lockdown and relaxation measures

To follow the SARS-CoV-2-detection rates after introducing more stringent lockdown measures in Switzerland in calendar week 12 (https://www.bag.admin.ch/bag/en/home/das-bag/aktuell/medienmitteilungen.msg-id-78454.html), we identified all NOPS from 12’363 symptomatic adults and children tested for SARS-CoV-2 from calendar week 14 to 24 (Follow-up cohort-1; **Table 1**) including 270 (2%) confirmed infections (**Figure 3A**). Despite continuing testing, the cumulative number of SARS-CoV-2 confirmed cases plateaued at approximately 1’300 by calendar week 18 (**Figure 3B**). After the initial rise until calendar week 12, the detection rates steadily declined from 13% to less than 1% by calendar week 19 (**Figure 3C**).

**Figure 3.**
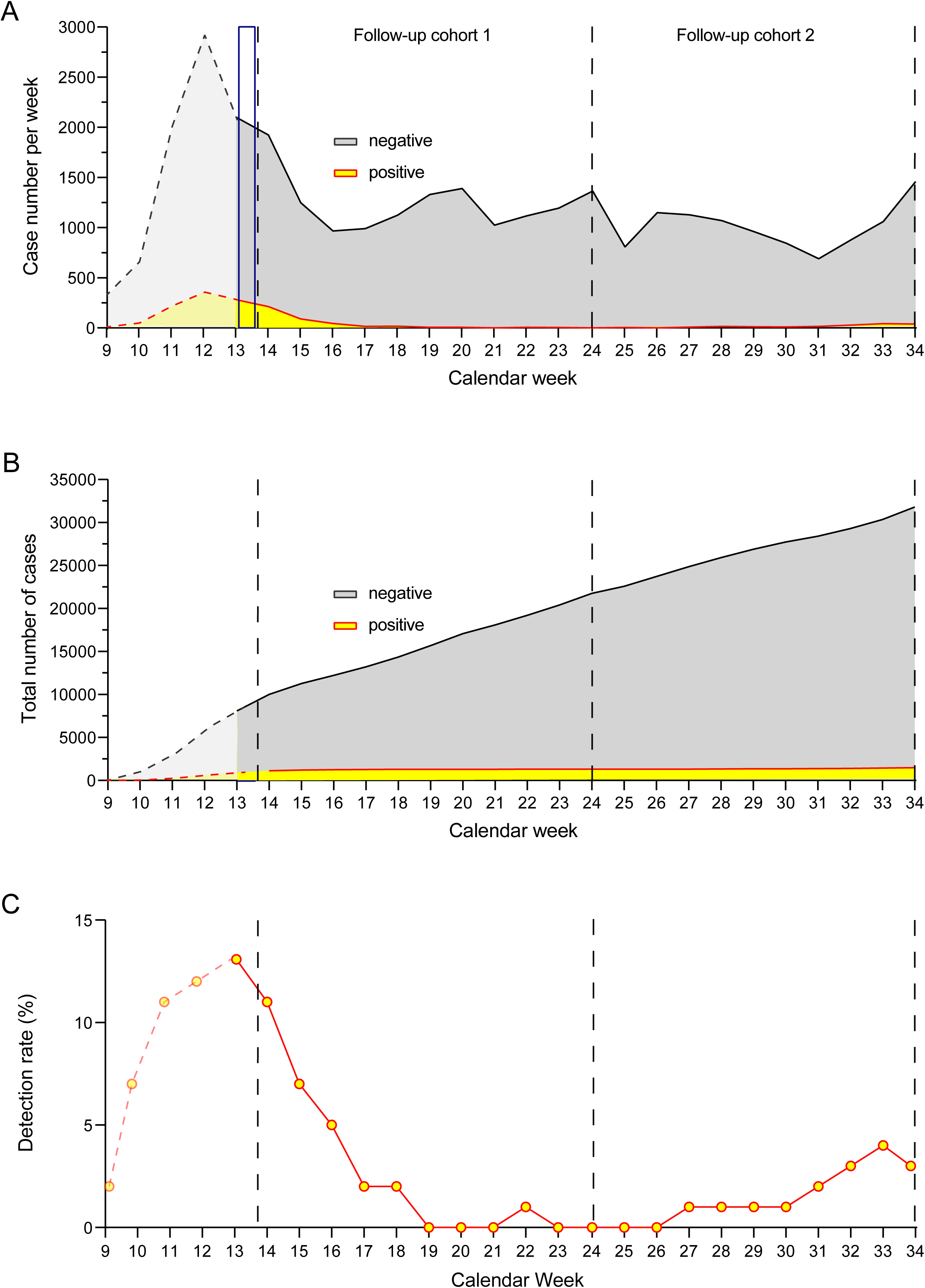
SARS-CoV-2 epidemiology in North-Western Switzerland from the beginning of the epidemic spread in February to the end of August 2020. The epidemiology and patients’ demographics of the first phase of the CoVID-19 pandemic from calendar week 9 to 13 has been recently reported [11]. The validation phase comparing the laboratory-developed Basel-S-gene, the commercial Roche-E-gene and the Roche-Cobas-Target1/Target2 assays in 1344 NOPS in calendar week 14 is indicated with a blue frame. The follow-up cohort-1 was analyzed from calendar week 14 to 24 (N=12’363), and follow-up cohort-2 from calendar week 24 to 34 (N=10’207). Patients’ demographics are displayed in Table 1. A. SARS-CoV-2 case number per week in symptomatic children and adults. B. Total SARS-CoV-2 detection from calendar week 9 to 34 in symptomatic children and adults. C. SARS-CoV-2 positivity rate from calendar week 9 to 34 in symptomatic children and adults.

During the next 10 week-period from calendar week 24 after relaxation of the lockdown measures to calendar week 34, another 10’207 patients were tested (Follow-up cohort-2; **Table 1**) including 176 (2%) confirmed SARS-CoV-2 infections. Most of these cases occurred after calendar week 26 showing an increase in the weekly SARS-CoV-2 detection rates to 4%. Comparing the patient demographics of the follow-up cohort-1 and -2 revealed a similar gender distribution, but patients with SARS-CoV-2 infections were significantly younger in the follow-up cohort-2 with a median age of 34 years (IQR 24 – 48) compared to 52 years (IQR 34 – 63) (p<0.001; **Table 1**). Also, the proportion of SARS-CoV-2 infected children had increased from 3% to 10% (p<0.01). The data indicated a changing epidemiological risk for the follow-up cohort-2 after lockdown measures had been relaxed in calendar week 24. As the declining prevalence impacts the pre-test probability and screening strategies, we examined the positive predictive value of SARS-CoV-2 detection using the automated testing platform. The positive predictive value of the Roche-Cobas-Target1 declined from 99.0% at detection rates of 15% seen before calendar 13 to 85.1% at rates 1% seen after calendar week 18 (**Supplementary Table S1**). However, the positive predictive values improved from 99.5% at 15% to 91.9% at 1% when combining the Roche-Cobas-Target1 and -Target2 results as a dual assay (**Supplementary Table S3**).

### Correlation and precision of SARS-CoV-2 RNA loads by different assays

Given the consistent association of low viral loads with discordant results and the natural course of SARS-CoV-2 infection, we systematically analyzed the precision using the Ct-values of the validation cohort. The overall Spearman correlation rS was significant for all assay comparisons ranging from 0.95 to 0.99 despite including the discordant results (**Figure 4; left panels**). When analyzing the precision of this quantitative correlation by Bland-Altmann analysis for the different assays (**Figure 4; middle panels**), higher Ct-values were needed for the Roche-E-gene compared to the Basel-S-gene assay with a mean bias of -2.8 cycles. When restricting the Bland-Altman analysis to the concordant samples, the mean difference was reduced by a Ct-value of approximately 1 (**Figure 4; right panels**).

**Figure 4.**
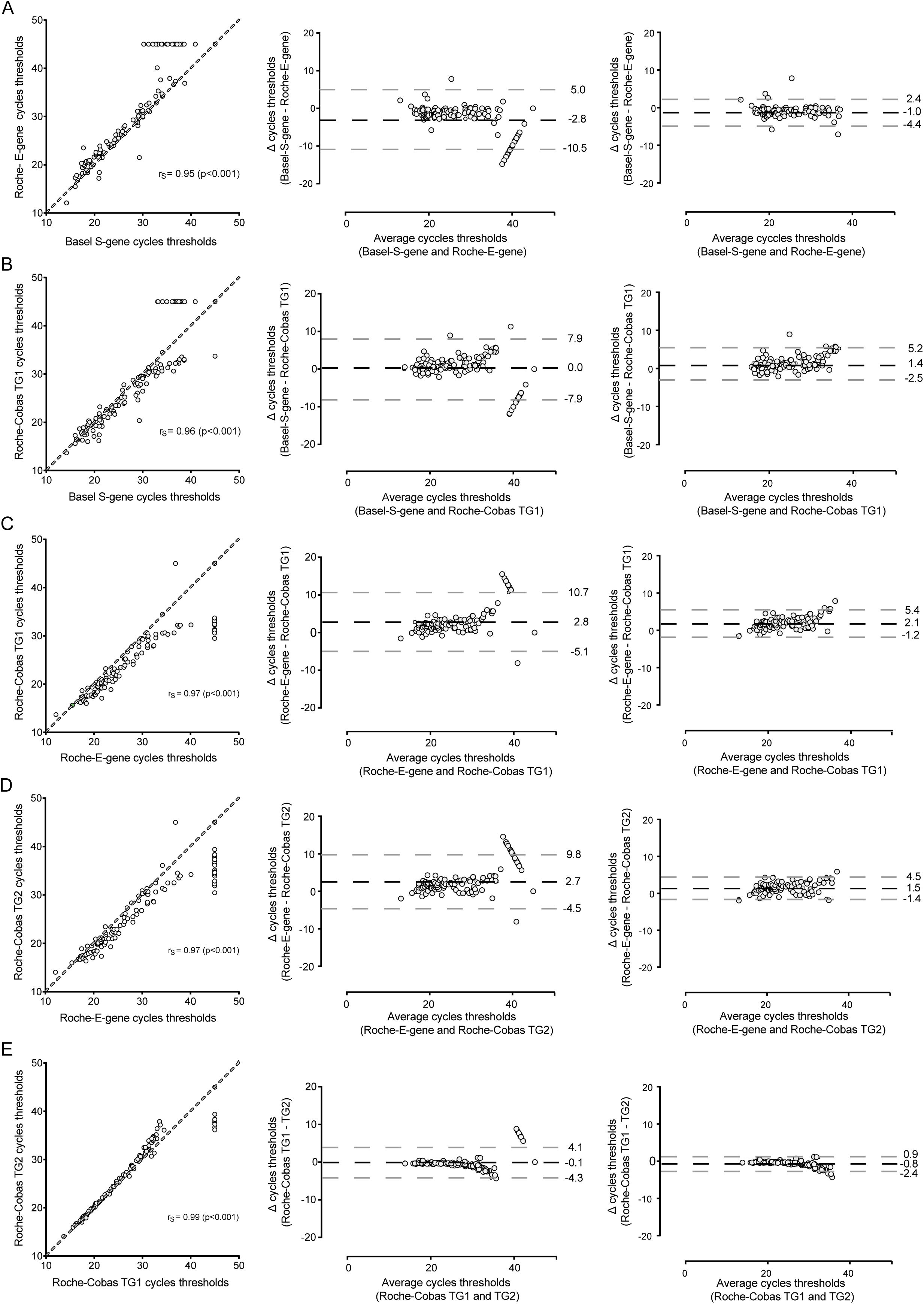
Spearman rank correlation and Bland-Altman analysis of 1344 prospectively compared NOPS using the laboratory-developed Basel-S-gene, the commercial Roche-E-gene and the Roche Cobas Target1/Target2 assays. A. Left panel: Spearman rank correlation of Basel-S-gene and Roche-E-gene cycling thresholds (dashed line indicates 100% agreement level). Middle panel: Bland-Altman analysis with a mean bias of −2.8 cycles, and 1.96 standard deviation of –10.5 and 5.0 cycles. Right panel: Bland-Altman analysis with a mean bias of −1.0 cycles, and 1.96 standard deviation of –4.4 and 2.4 cycles. B. Left panel: Spearman rank correlation of Basel-S-gene and Roche-Cobas-Target1 cycling thresholds (dashed line indicates 100% agreement level). Middle panel: Bland-Altman analysis with a mean bias of 0.0 cycles, and 1.96 standard deviation of –7.9 and 7.9 cycles. Right panel: Bland-Altman analysis with a mean bias of 1.4 cycles, and 1.96 standard deviation of –2.5 and 5.2 cycles. C. Left panel: Spearman rank correlation of Roche-E-gene and Roche-Cobas-Target1 cycling thresholds (dashed line indicates 100% agreement level). Middle panel: Bland-Altman analysis with a mean bias of 2.8 cycles, and 1.96 standard deviation of –5.1 and 10.7 cycles. Right panel: Bland-Altman analysis with a mean bias of 2.1 cycles, and 1.96 standard deviation of –1.2 and 5.4 cycles. D. Left panel: Spearman rank correlation of Roche-E-gene and Roche-Cobas-Target2 cycling thresholds (dashed line indicates 100% agreement level). Middle panel: Bland-Altman analysis with a mean bias of 2.7 cycles, and 1.96 standard deviation of –4.5 and 9.8 cycles. Right panel: Bland-Altman analysis with a mean bias of 1.5 cycles, and 1.96 standard deviation of –1.4 and 4.5 cycles. E. Left panel: Spearman rank correlation of Roche-Cobas-Target1 and Roche-Cobas-Target2 cycling thresholds (dashed line indicates 100% agreement level). Middle panel: Bland-Altman analysis with a mean bias of -0.1 cycles, and 1.96 standard deviation of –4.3 and 4.1 cycles. Right panel: Bland-Altman analysis with a mean bias of -0.8 cycles, and 1.96 standard deviation of –2.4 and 0.9 cycles.

Comparing the Basel-S-gene and Roche-Cobas-Target1 revealed a mean difference of 0.0 cycles for all comparisons, and +1.4 cycles for the concordant positives corresponding to 0.42 log10 copies/mL. Together, the data indicated a high quantitative correlation of the manual and the automated SARS-CoV-2 detection assays with good precision permitting to take the Basel-S-gene and Roche-Cobas-Target1 for further study.

To examine the SARS-CoV-2 loads in follow-up NOPS of patients with confirmed SARS-CoV-2 diagnosis, we analyzed viral loads in 936 follow-up NOPS submitted from 261 patients (**Supplementary Table S4**). The median age of the patients was 60 years, 60% were males, and two-thirds were hospitalized including 21% of patients on the intensive care unit. Although some variability was seen especially within the first 5 days, the overall results showed that SARS-CoV-2 loads in NOPS significantly declined with increasing time after the initial diagnosis (**Figure 5A**). At 10 days, SARS-CoV-2 loads were approximately 3 log10 copies/mL lower than at diagnosis. Twenty days after the initial diagnosis, persisting SARS-CoV-2 loads mostly below 10’000 c/mL. Among 158 patients with documented viral clearance in NOPS, the median time to undetectable was 14 days (IQR: 5 – 21; data not shown). Analysing 79 patients with high sampling density of three and more NOPS similarly showed an approximately 3 log10 decline at 10 days post-diagnosis and a median time to undetectable of 14 days (IQR: 9 – 20; **Supplementary Figure S2**).

**Figure 5.**
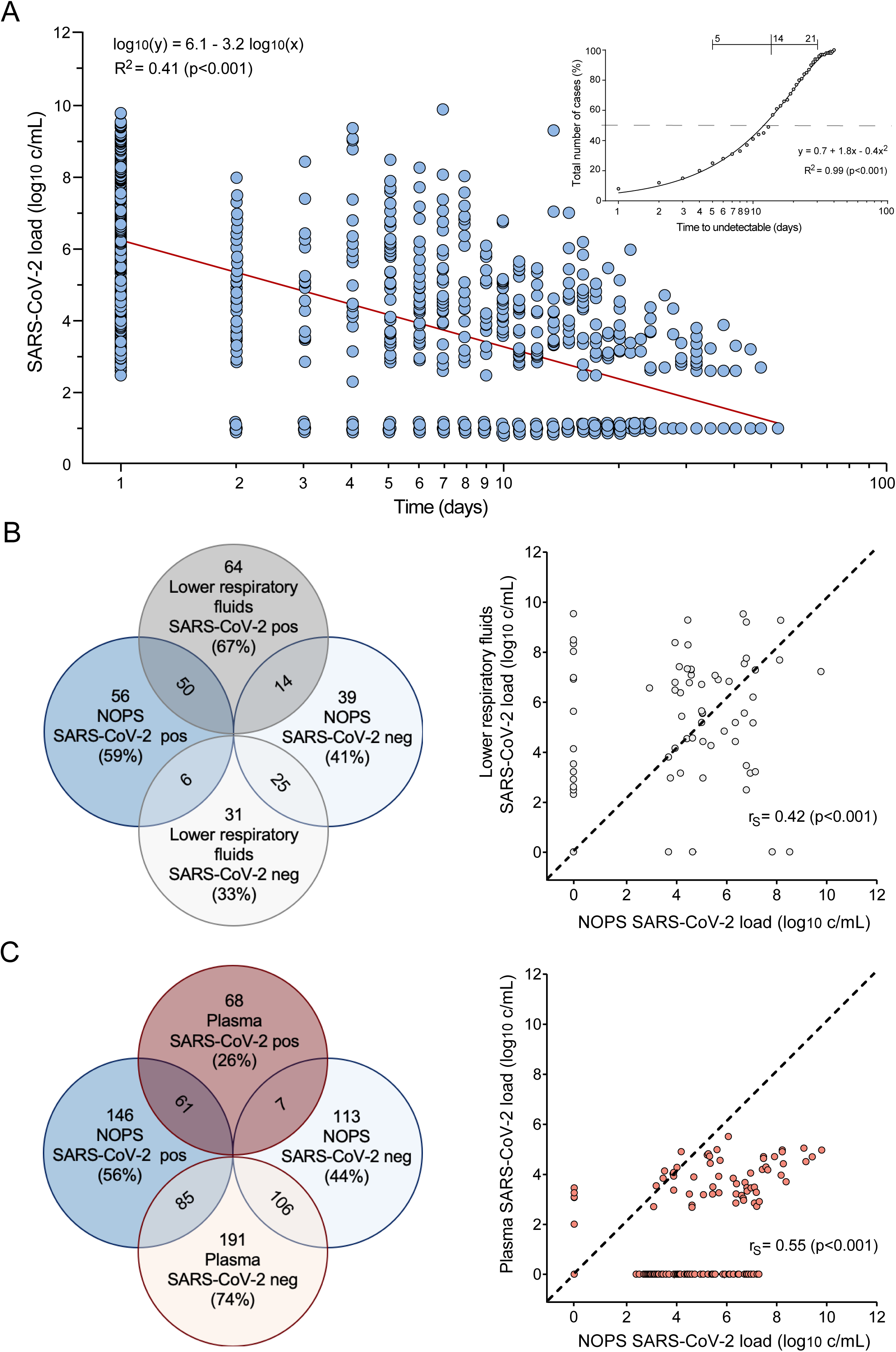
Quantification of SARS-CoV-2 RNA loads in NOPS at diagnosis and follow-up, in time-matched lower respiratory fluids and in plasma samples. A. SARS-CoV-2 RNA loads in NOPS at diagnosis and at different times of follow-up consisting of 936 samples submitted from 261 patients. SARS-CoV-2 loads below the limit of detection are arbitrarily set to 1 log10 c/mL. Red line shows the regression analysis of log transformed SARS-CoV-2 loads. The inset summarizes the events over time of SARS-CoV-2-RNA-positive (red); undetectable (blue); and positive without follow-up testing (grey). B. VENN diagram of SARS-CoV-2 detection in parallel sampled NOPS and lower respiratory fluids using Basel-S-gene RT-QNAT (N=95; left panel). Spearman rank correlation of SARS-CoV-2 loads in NOPS and lower respiratory fluids (dashed line indicates 100% agreement level; right panel). C. VENN diagram of SARS-CoV-2 detection in parallel sampled NOPS and plasmas using Basel-S-gene RT-QNAT (N=259; left panel). Spearman rank correlation of SARS-CoV-2 loads in NOPS and plasmas (dashed line indicates 100% agreement level; right panel). The respective patients’ demographics are summarized in Supplementary Table S4.

To compare the quantitative relationship of SARS-CopV-2 loads in NOPS and lower respiratory fluid samples, we identified 36 CoVID-19 patients providing 95 time-matched NOPS and lower respiratory fluid samples taken within <7 days (**Supplementary Table S4**). The median age was 60 years, 97% were hospitalized and 75% on the intensive care unit. Spearman analysis revealed a weak but significant quantitative correlation (rS=0.42; p<0.001; **Figure 5B**). In 14 pairs, SARS-CoV-2 was still detectable in lower respiratory fluids, having median viral RNA loads of 4.9 log10 c/mL.

To investigate the quantitative relationship of SARS-CopV-2 loads in NOPS and plasma viral loads, we identified 129 CoVID-19 patients providing 259 NOPS and time-matched plasma samples (**Supplementary Table S4**). In 61 of 68 NOPS-positive pairs, SARS-CoV-2 was also detectable in plasma, giving rise to a weak but significant quantitative correlation (Spearman, rS=0.55; p<0.001; **Figure 5C**). In 7 pairs, SARS-CoV-2 was detectable in plasma at low viral loads of 3 log10 c/mL, but no longer in the time-matched NOPS.

Taken together, the results supported the notion that quantification of viral loads in different analytes provided a limited, but clinically and epidemiologically relevant first insight of SARS-CoV-2 replication dynamics over time and the role of different anatomic sampling compartments.

## Discussion

Reliable SARS-CoV-2 detection is key to the clinical and the epidemiological assessment during the current CoVID-19 pandemic. Both goals are critically dependent on the performance of diagnostic assays, which again are impacted by the prevalence and dynamics following effective lockdown measures and their relaxation. There are only limited real-life data available addressing these diagnostic and epidemiologic challenges in a comprehensive manner. In this study, we report the qualitative and quantitative analysis of SARS-CoV-2 QNAT assays for the diagnosis and in clinically relevant settings of patient follow-up and describe the impact following lockdown and relaxation in Northwestern Switzerland. Our prospective cross-validation of 1344 NOPS demonstrates that a fully automated dual-target assay has a high positive and negative predictive value of 99.5% and 98.7%, respectively, at a prevalence of 15% for a qualitative result, which changes to 91.9% and 99.9% at rates around 1%. Expectedly, single-target assays are more dramatically affected. Thus, this real-life setting indicates the need for confirmatory information in up to 8% of clinical cases and point to critical limitations of large-scale screening of the general population and impact the validity of sample pooling endeavors. Both approaches are discussed for assessing periods of effective containment as well as for testing populations with difficult to define SARS-CoV-2 exposure risk such as returning travelers.

In the clinical setting of patients presenting to our center, SARS-CoV-2 detection steadily declined from 9% in the validation cohort close to the peak of the first pandemic wave to an overall rate of 2% in both of our two follow-up cohorts, each covering more than 10’000 patients over 10 weeks. The epidemiologic dynamics were captured by weekly detection rates, which steadily declined to <1% and then flared up to 4%, both at approximately 6 weeks following lockdown and relaxation measures, respectively. Although we cannot define the key factors for either development, the demographics reveal a significantly younger median age of only 34 years of SARS-CoV-2 diagnosis in the follow-up cohort-2 compared to approximately 50 years in the validation and follow-up cohort-1. Although the proportion of children testing positive for SARS-CoV-2 had also significantly increased from 3% to 10%, the interquartile age range of follow-up cohort-2 indicated that the recent SARS-CoV-2 flare was mostly due to younger adults between 24 and 48 years of age. These data suggest that behavioral factors were underlying these developments, which might reflect older populations remaining more adherent to (self-)protective measures, while younger adults accepted increased exposure risks such traveling and gathering in larger groups during this period of summer holidays and school closure in Switzerland.

Our detailed study of SARS-CoV-2 quantification added an independent layer of confirmatory evidence in low prevalence settings. SARS-CoV-2 positive NOPS showed high median viral loads of 10 million copies per milliliter of sample (IQR 130’000 - 150 million) in our validation cohort and were equally high in patients diagnosed during the pandemic rise [11] and during off-peak times with detection rates of 2%. Although no commutable validated calibrator is available, highly significant quantitative correlations were observed for all SARS-CoV-2 assays analyzed. The detailed Bland-Altman analysis enumerated the precision in Ct-values when comparing the different assays. Specifically, the automated Roche-Cobas-Target1 and the Basel-S-gene could be used interchangeably as mean differences of Ct-values ranged from 0.0 to 1.4; hence, being within the difference of 0.3 – 0.5 log_10_ commonly accepted in QNAT comparisons [17, 18]. Given the differences in design, targets, extraction, and hard ware between our laboratory-developed and commercial assays, these strong correlations provided the technical basis for considering high SARS-CoV-2 loads in NOPS in support of the diagnosis.

Conversely, discordant results between the different assays occurred in <3% and were characterized by significantly lower SARS-CoV-2-loads close to the limit of detection. In half of these cases, SARS-CoV-2 infection could be independently confirmed, again having low viral loads. Among the 1220 NOPS having undetectable SARS-CoV-2 RNA by both, Roche-Cobas-Target1 and -Target2, only 9 NOPS (0.7%) were independently confirmed as being low SARS-CoV-2-positive, in line with the high negative predictive values. Although details of the commercial assays were not available, our analysis of more than 25’000 complete SARS-CoV-2 genomes provided no evidence for relevant target sequence changes of the laboratory-developed assays. Together, the data support the view that high viral loads confirmed the diagnosis, whereas low viral loads close to the limit of detection were associated with discordant or potentially missed diagnosis, for which confirmatory testing should be considered.

Quantifying viral RNA in NOPS provided independent evidence that SARS-CoV-2 loads decreased by approximately 3 log10 copies per milliliter NOPS at 10 days after diagnosis. This observation is notable for several reasons: First, the data indicate a fairly rapid decline of viral replication in the upper respiratory tract in the absence of effective antiviral treatment. Second, the window for confirmatory NOPS testing is rather short for diagnosing patients having low viral loads or ambiguous results. Third, in symptomatic patients with undetectable SARS-CoV-2 results in NOPS, the potential diagnostic utility of SARS-CoV-2 in lower respiratory fluids or in plasma should be considered [19].

The comparison of time-matched samples from the upper and lower respiratory tract revealed a weak, but significant correlation. However, in 14/95 pairs, SARS-CoV-2 was below the limit of detection in NOPS, but still detectable in lower respiratory fluids with median viral loads close to 1 million copies per milliliter. A similarly weak correlation was observed for NOPS and plasma viral loads, and although in 7 cases, SARS-CoV-2 RNA was detectable in plasma but not in NOPS. Taken together, individual confirmatory and quantitative follow-up testing should be considered within few days, submitting lower respiratory fluids from symptomatic patients with SARS-CoV-2-negative NOPS. Other studies have reported similar rule-of thumb dynamics of SARS-CoV-2-loads in upper respiratory fluids of symptomatic, pre-symptomatic and more recently in asymptomatic infected individuals, whereby disease severity, older age and immunodeficiency have been associated with prolonged shedding in some but not all studies [20-22].

## Limitations

Limitations of our study are the observational character of this data reflecting the real-life setting and the fact that we have no comprehensive data regarding duration and severity of symptoms of the approximately 24’000 patients. However, most patients diagnosed in the validation and the follow-up cohorts presented to our outpatient clinic suggesting an illness in line with the current recommendations but no need for immediate hospitalization. A recent study reported that medical attention was sought on average 3 to 5 days after symptoms onset similar to other influenza-like illnesses [19]. Conversely, most of our 261 patients providing follow-up NOPS were hospitalized and in their sixth age decade associated with more severe disease including the need for intensive care. Although we cannot exclude that the median time to undetectable NOPS viral load may be shorter in a- or oligosymptomatic infections than in our patients, the 99.9% decline within 10 days in all 261 patients or in a subgroup of 79 patients with very high sampling density and their median time to undetectable SARS-CoV-2 in NOPS at 14 days (IQR 9 – 20) after diagnosis provides some orientation for confirmatory diagnostics and infection control measures.

## Conclusion

This prospective cross-validation of 1344 NOPS demonstrates that a fully automated dual-target assay has a high positive and negative predictive value of 99.5% and 98.7%, respectively, at a prevalence of 15% for a qualitative result, which changes to 91.9% and 99.9% at rates around 1%. The qualitative concordance of the two automated and the two manual assays is 97%, while the discordant results are due to limiting SARS-CoV-2 RNA loads in the NOPS. High quantitative correlations were observed between all assays, permitting to consider SARS-CoV-2 RNA loads in follow-up testing as well as in other biological fluids such as specimens from the lower respiratory tract. Given the rapid decline of SARS-CoV-2 loads of 3 log_10_ copies/mL at 10 days after initial diagnosis, follow-up testing of NOPS should be considered early after initial testing, preferably with less than 5 days. In symptomatic patients with an epidemiologic link and SARS-CoV-2-undetectable NOPS, testing of from lower respiratory fluids may be considered.

## Data Availability

All data are available at a secure server accessed only by the research team, but restrictions apply to guarantee pseudonymized patient identity.

## Acknowledgements

We thank the biomedical analysts of the Clinical Virology, Laboratory Medicine, University Hospital Basel, Basel, Switzerland, for expert help and assistance.

## Declarations

### Author contributions

KL and HHH designed the study, validated results, extracted data, performed analyses, reviewed data and wrote the manuscript; RG, KKS, KN, TR, KR, and AE validated results,

analyzed data and contributed to writing to the manuscript; JB, RB, CHN, NK, STS, AFW, HP, MS, DS, MT, SB, MO, MB evaluated patients, submitted samples, reviewed data and contributed to writing the manuscript.

## Supplementary Material

This supplementary material has supporting information alongside the article [*Epidemiology and precision of SARS-CoV-2 detection following lockdown and relaxation measures*], on behalf of the authors.

## Supplementary Tables

**Table S1.** SARS-CoV-2 rates and Roche-cobas®6800 platform test characteristics.

| Cobas Target 1 | COVID-19 prevalence | 1% | 2.5% | 5% | 7.5% | 10% | 12.5% | 15% | 17.5% | 20% |
| --- | --- | --- | --- | --- | --- | --- | --- | --- | --- | --- |
|  | Sensitivity | 92.3% | 92.3% | 92.3% | 92.3% | 92.3% | 92.3% | 92.3% | 92.3% | 92.3% |
|  | Specificity | 99.8% | 99.8% | 99.8% | 99.8% | 99.8% | 99.8% | 99.8% | 99.8% | 99.8% |
|  | False Positive Rate | 0.16% | 0.16% | 0.16% | 0.16% | 0.16% | 0.16% | 0.16% | 0.16% | 0.16% |
|  | False Negative rate | 7.7% | 7.7% | 7.7% | 7.7% | 7.7% | 7.7% | 7.7% | 7.7% | 7.7% |
|  | Positive Likelihood Ratio | 566.3 | 566.3 | 566.3 | 566.3 | 566.3 | 566.3 | 566.3 | 566.3 | 566.3 |
|  | Negative Likelihood Ratio | 0.08 | 0.08 | 0.08 | 0.08 | 0.08 | 0.08 | 0.08 | 0.08 | 0.08 |
|  | Positive Predictive Value* | 85.1% | 93.6% | 96.8% | 97.9% | 98.4% | 98.8% | 99.0% | 99.2% | 99.3% |
|  | Negative Predictive Value* | 99.9% | 99.8% | 99.6% | 99.4% | 99.2% | 98.9% | 98.7% | 98.4% | 98.1% |
|  | Accuracy* | 99.8% | 99.7% | 99.5% | 99.3% | 99.1% | 98.9% | 98.7% | 98.5% | 98.3% |

| Cobas Target 2 | COVID-19 prevalence | 1% | 2.5% | 5% | 7.5% | 10% | 12.5% | 15% | 17.5% | 20% |
| --- | --- | --- | --- | --- | --- | --- | --- | --- | --- | --- |
|  | Sensitivity | 92.3% | 92.3% | 92.3% | 92.3% | 92.3% | 92.3% | 92.3% | 92.3% | 92.3% |
|  | Specificity | 99.2% | 99.2% | 99.2% | 99.2% | 99.2% | 99.2% | 99.2% | 99.2% | 99.2% |
|  | False Positive Rate | 0.82% | 0.82% | 0.82% | 0.82% | 0.82% | 0.82% | 0.82% | 0.82% | 0.82% |
|  | False Negative rate | 7.7% | 7.7% | 7.7% | 7.7% | 7.7% | 7.7% | 7.7% | 7.7% | 7.7% |
|  | Positive Likelihood Ratio | 113.3 | 113.3 | 113.3 | 113.3 | 113.3 | 113.3 | 113.3 | 113.3 | 113.3 |
|  | Negative Likelihood Ratio | 0.08 | 0.08 | 0.08 | 0.08 | 0.08 | 0.08 | 0.08 | 0.08 | 0.08 |
|  | Positive Predictive Value* | 53.4% | 74.4% | 85.6% | 90.2% | 92.6% | 94.2% | 95.2% | 96.0% | 96.6% |
|  | Negative Predictive Value* | 99.9% | 99.8% | 99.6% | 99.4% | 99.2% | 98.9% | 98.7% | 98.4% | 98.1% |
|  | Accuracy* | 99.1% | 99.0% | 98.8% | 98.7% | 98.5% | 98.3% | 98.2% | 98.0% | 97.8% |

\*depend on SARS-CoV-2 detection rates

**Table S2.** NOPS from patients with human coronavirus (HCoV) infections other than SARS-CoV-2 were used to assess the specificity of the different assays.

| Patient nr. | Multiplex NAT | Roche-Cobas-Target1<sup>a</sup> | Roche-Cobas-Target2<sup>b</sup> | Basel-S-gene | Roche-E-Gene | Basel-ORF8-gene |
| --- | --- | --- | --- | --- | --- | --- |
| 1 | HCoV HKU1 | - | - | - | - | - |
| 2 | HCoV HKU1 | - | - | - | - | - |
| 3 | HCoV HKU1 | - | - | - | - | - |
| 4 | HCoV HKU1 | - | - | - | - | - |
| 5 | HCoV HKU1 | - | - | - | - | - |
| 6 | HCoV NL63 | - | - | - | - | - |
| 7 | HCoV NL63 | - | - | - | - | - |
| 8 | HCoV NL63 | - | - | - | - | - |
| 9 | HCoV NL63 | - | - | - | - | - |
| 10 | HCoV NL63 | - | - | - | - | - |
| 11 | HCoV OC43 | - | - | - | - | - |
| 12 | HCoV OC43 | - | - | - | - | - |
| 13 | HCoV OC43 | - | - | - | - | - |
| 14 | HCoV OC43 | - | - | - | - | - |
| 15 | HCoV OC43 | - | - | - | - | - |
| 16 | HCoV 229E | - | - | - | - | - |
| 17 | HCoV 229E | - | - | - | - | - |
| 18 | HCoV 229E | - | - | - | - | - |
| 19 | HCoV 229E | - | - | - | - | - |
| 20 | HCoV 229E | - | - | - | - | - |

E-gene, envelope gene; HCoV, human coronavirus; SARS-CoV-2, severe acute respiratory syndrome coronavirus 2; S-gene, spike glycoprotein gene; ORF8-gene, open reading frame 8 gene
<sup>a</sup> targets the open reading frame 1 non-structural region that is unique to SARS-CoV-2
<sup>b</sup> targets a conserved region in the structural protein envelope gene present in all Sarbecoviruses

**Table S3.** SARS-CoV-2 rates and Roche-cobas®6800 test characteristics of the combined Target1 and Target2.

| **Cobas Target 1 / Target 2** | **COVID-19 prevalence** | **1%** | **2.5%** | **5%** | **7.5%** | **10%** | **12.5%** | **15%** | **17.5%** | **20%** |
| --- | --- | --- | --- | --- | --- | --- | --- | --- | --- | --- |
|  | Sensitivity | 92.3% | 92.3% | 92.3% | 92.3% | 92.3% | 92.3% | 92.3% | 92.3% | 92.3% |
|  | Specificity | 99.9% | 99.9% | 99.9% | 99.9% | 99.9% | 99.9% | 99.9% | 99.9% | 99.9% |
|  | False Positive Rate | 0.08% | 0.08% | 0.08% | 0.08% | 0.08% | 0.08% | 0.08% | 0.08% | 0.08% |
|  | False Negative rate | 7.7% | 7.7% | 7.7% | 7.7% | 7.7% | 7.7% | 7.7% | 7.7% | 7.7% |
|  | Positive Likelihood Ratio | 1132.6 | 1132.6 | 1132.6 | 1132.6 | 1132.6 | 1132.6 | 1132.6 | 1132.6 | 1132.6 |
|  | Negative Likelihood Ratio | 0.08 | 0.08 | 0.08 | 0.08 | 0.08 | 0.08 | 0.08 | 0.08 | 0.08 |
|  | Positive Predictive Value* | 92.0% | 96.7% | 98.4% | 98.9% | 99.2% | 99.4% | 99.5% | 99.6% | 99.7% |
|  | Negative Predictive Value* | 99.9% | 99.8% | 99.6% | 99.4% | 99.2% | 98.9% | 98.7% | 98.4% | 98.1% |
|  | Accuracy* | 99.8% | 99.7% | 99.5% | 99.4% | 99.2% | 99.0% | 98.8% | 98.6% | 98.4% |

\*depend on SARS-CoV-2 detection rates

**Table S4.**
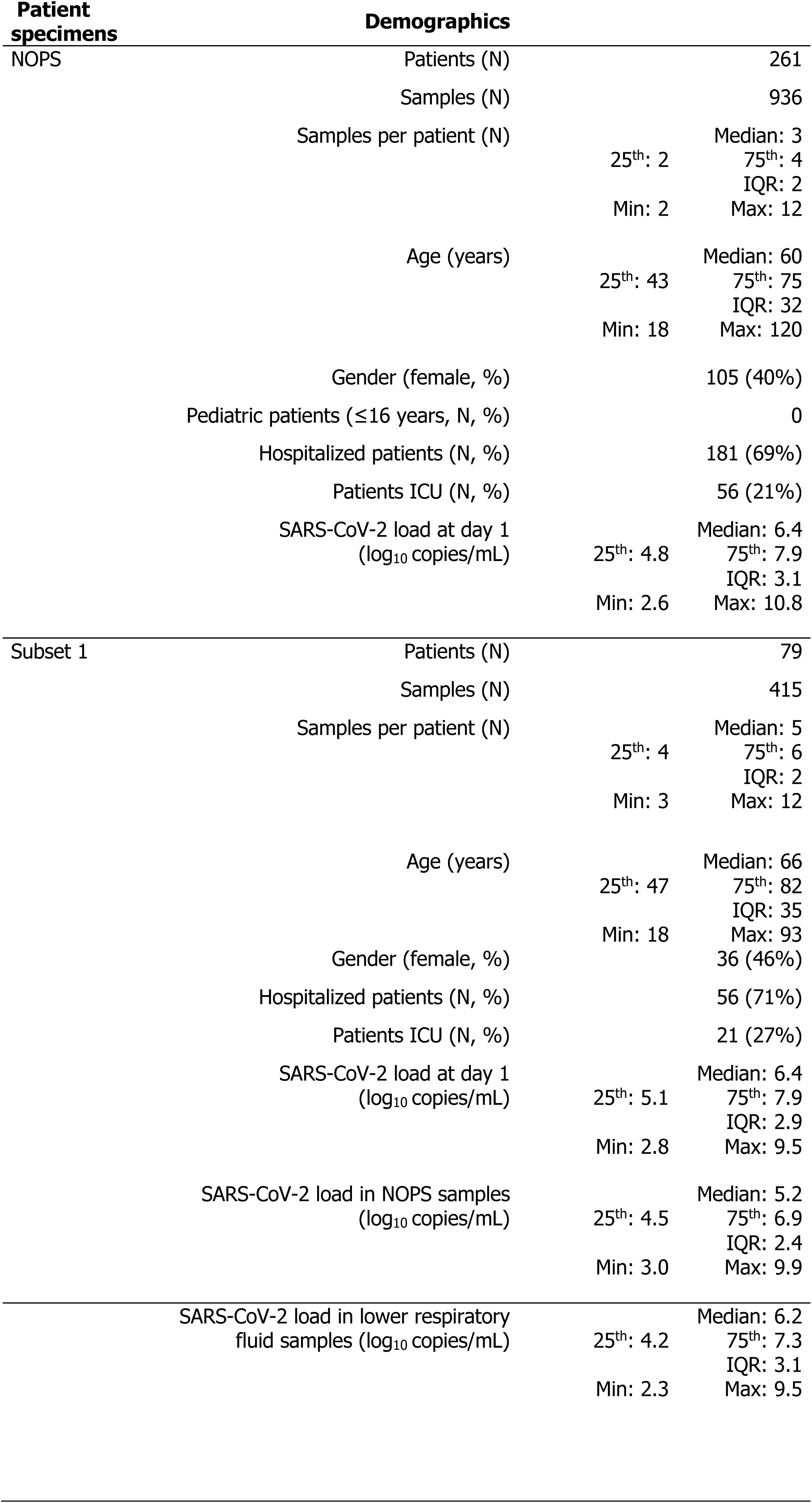
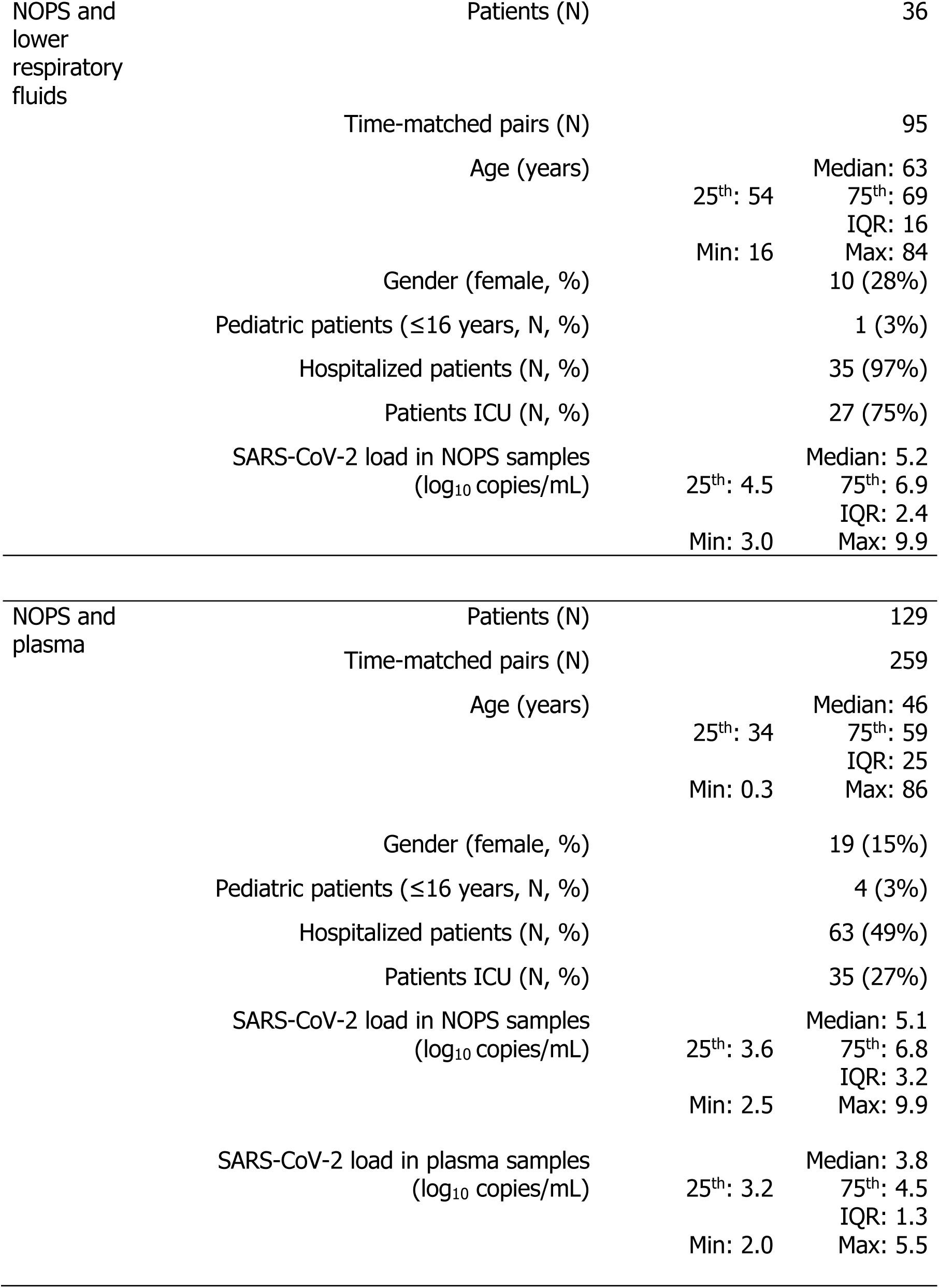
Quantification of SARS-CoV-2 in NOPS and lower respiratory fluids and plasma samples, respectively of COVID-19 patients using Basel-S-gene RT-QNAT.

## Supplementary Figures

**Figure S1.**
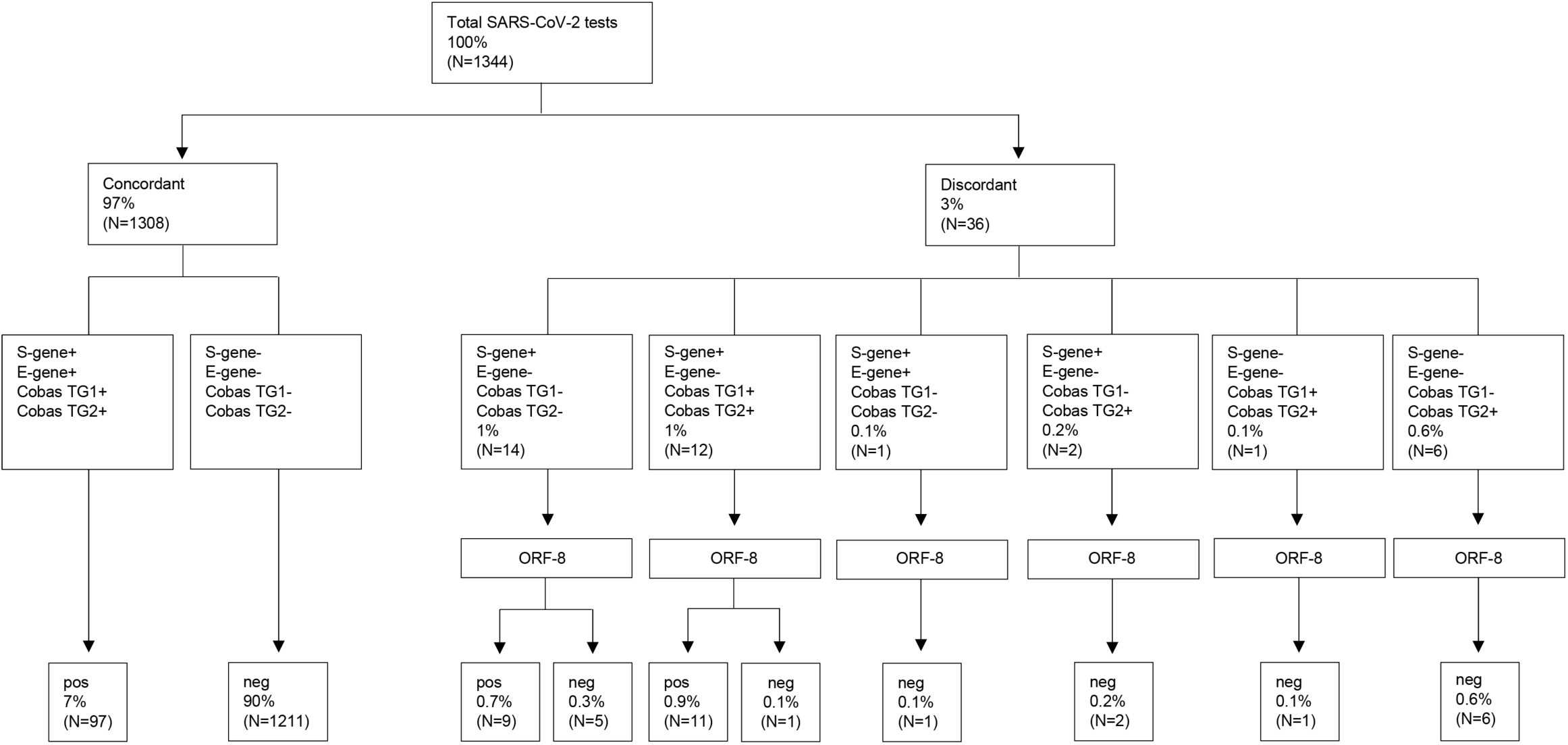
SARS-CoV-2 testing flowchart. NOPS were prospectively tested in parallel with the laboratory-developed Basel-S-gene, the commercial Roche-E-gene and the Roche-Cobas-Target1/Target2 assays (n=1344). Samples with discordant results were subsequently tested with the laboratory-developed Basel-ORF8-gene RT-QNAT. pos, positive; neg, negative.

**Figure S2.**
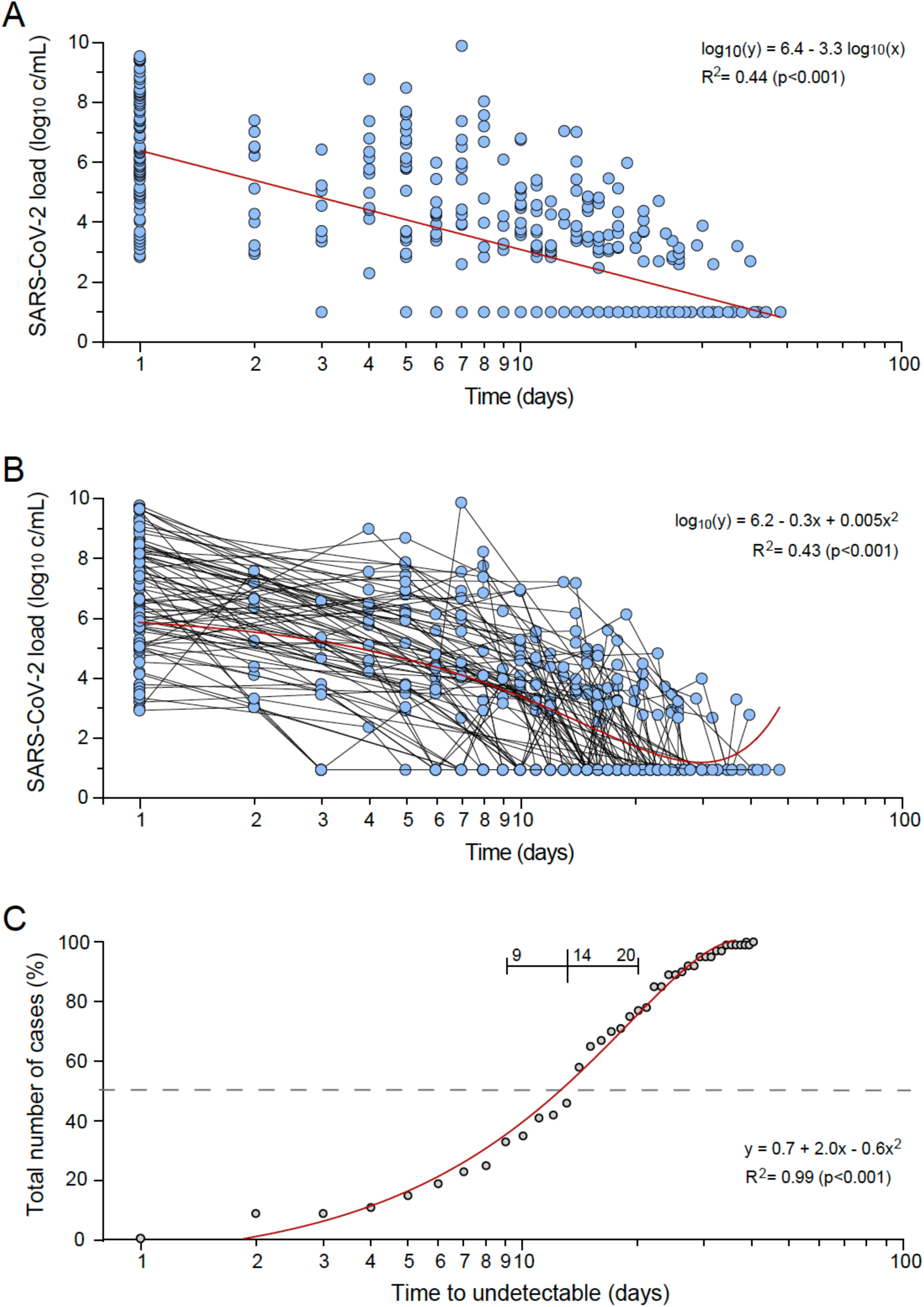
Time to undetectable SARS-CoV-2 in NOPS of patients with frequent NOPS sampling after diagnosis. A subset 1 of 79 patients of the 261 patients presented in Figure 5A and supplementary Table 4 were identified fulfilling the following three criteria i) at least three and more NOPS samples analyzed ii) at least one positive sample confirming the initial SARS-CoV-2 diagnosis; iii) at least one NOPS sample analyzed <14 days after SARS-CoV-2 diagnosis. A. SARS-CoV-2 RNA load in NOPS over time after diagnosis using linear curve fitting; B. SARS-CoV-2 RNA load in NOPS over time after diagnosis using polyonomial curve fitting accounting for rebound detection at low viral loads; C. Time to first undetectable SARS-CoV-2 RNA in NOPS (line cross: median time and interquartile range).

